# The Effect of Temperature on Covid-19 Confirmed Cases: Evidence from US Counties

**DOI:** 10.1101/2021.01.08.21249449

**Authors:** Hamid NoghaniBehambari, Mahmoud Salari, Farzaneh Noghani, Nahid Tavassoli

## Abstract

This paper studies the effect of air temperature on the transmission of COVID-19 in the U.S. using daily observations across counties. This study uses various ordinary least squares (OLS) models with a comprehensive set of fixed effects to overcome unobserved heterogeneity issues across counties as well as the generalized method of moments (GMM) estimators as dynamic models to address endogeneity issue. Our main results indicate that an increase of one degree in temperature is associated with a reduction of 0.041 cases per 100,000 population at the county-level. We run several robustness tests and all the models confirm the impact of temperature on COVID-19 confirmed new cases. These results help policymakers and economists in optimizing decisions and investments to reduce COVID- 19 new cases.

**JEL Codes:** I10; Q51; Q54; H12

## 1. Introduction

The world has been faced with the pandemic issue, COVID-19, in 2020 which has impacted all countries and their economies. This pandemic reminds us of the last pandemic of the world which was the 1918 influenza pandemic, started in London, United Kingdom. The number of people death associated with COVID-19 is much higher than previous coronaviruses (i.e. SARS-CoV and MERS-CoV) so far and still goes up with a huge impact on economies(Bogoch et al., 2020; Q. Lin et al., 2020). The COVID-19 pandemic has impacted both demand and supply of commodities across the countries in both short-term and long-term. In the short-term, as many regions (countries and states) adopt strict regulations, their economies slow down significantly^4^. The long-term impacts of this pandemic are a sharp rise in unemployment for individuals and bankruptcy and a hard time to survive for many businesses(Goodell, 2020; Zhang, Hu, and Ji, 2020).

Based on the official reports, the U.S. has the highest number of infected people (around 30%) by COVID-19 around the world. The amount of money that U.S. imposed on the economy to support the economy and prevent future recession regarding COVID-19 is more than many countries’ GDP so far and expects to go higher in near future which would bring more debt for the Federal Reserve has recently announced that interest rates would drop to a range of 0%^5^ to 0.25% and buy at least $700 billion in government and mortgage-related bonds as a part of their emergency actions to protect the economy regarding the economic impacts of coronavirus (Long, 2020). The U.S. House passed a $3 trillion coronavirus relief bill on May 2020 (Bellware et al., 2020) and many economists believe that the U.S. government has committed more than $6 trillion on the economy during this pandemic (Dam, 2020). Thus, understanding COVID-19 and estimating the potential factors that may influence its spread is very important for policymakers and economists.

This study uses an extensive daily panel data sets and applying both static and dynamic analysis to find the potential link between the level of temperatures across counties and the number of COVID-19 confirmed new cases. Using a comprehensive set of fixed effects, we find a negative relationship between lagged levels of temperature and the new confirmed cases. An increase of 10 °F in temperature of the last 3-14 days reduces the incidence of new confirmed cases by 0.45 counts per 100,000 population, a reduction of roughly 25% from the mean of daily confirmed cases at the county level.

The contribution of this study to the literature is twofold. First, this is the first county-level study clarifying the relationship between temperature and COVID-19 new cases. Second, this study applies several different approaches including static models and dynamic models that allow us to control for endogeneity and omitted variable biases.

The rest of the study is organized in the following way: Section 2 provides a literature review; Section 3 shows data and sample selection strategy; Section 4 demonstrates the empirical strategy; Section 5 reports the results; Section 6 presents discussion and conclusion.

## 2. Literature Review

Coronaviruses are a family of viruses that can result in different types of illnesses such as severe acute respiratory syndrome. A novel coronavirus outbreak started in late 2019 in China and has since spread at a rapid pace throughout the world. COVID-19 is the respiratory infectious disease caused by the novel coronavirus. Within a month of the first reported case in China, horrifying numbers of infections and deaths followed in countries across the globe. The first confirmed case of COVID-19 in the US was reported on January 21, 2020, in the state of Washington. Since then, this disease has spread to every state in the US and infected more than one million people (CDC, 2020).

Researchers have found that COVID-19 is mainly transmitted through respiratory droplets from infected people with symptoms to others and by direct contact with infected people, contaminated surfaces, and contaminated objects (Liu et al., 2020; Ong et al., 2020). Research on the preceding types of the novel coronavirus shows that several factors can affect their transmission. These factors include population density, quality of medical care, ambient temperature, and humidity (Barreca and Shimshack, 2012; Casanova, Jeon, Rutala, Weber, and Sobsey, 2010; Dalziel et al., 2018; Lowen, Mubareka, Steel, and Palese, 2007; Noghani and Noghanibehambari, 2019).

Among the factors influencing the transmission and survival of coronaviruses, the air temperature has received considerable attention (Otter et al., 2016). Other than the fact that the virus half-life is increased as temperature decreases (Lowen and Steel, 2014), ambient temperature can affect the transmission and survival of coronavirus, and consequently the number of infected people, through two main mechanisms. The first mechanism is that as temperature decreases, the spread of the virus in the nasal mucosa is improved (Lowen et al., 2007). When the temperature decreases, cooler air is breathed and thus the nasal mucosa is cooled. Consequently, the viscosity of the mucous layer increases and the frequency of cilia beats decreases. These conditions will slow down a mucociliary clearance and help speed the process of spreading the virus into the respiratory tract (Eccles, 2002). In other words, as temperature decreases, the immunity to the virus infection decreases(Eccles, 2002; Lowen and Steel, 2014). Furthermore, when the nasal mucosa is cooled, the virus that has entered the airways will persist there longer (i.e., its half-life is increased) due to the decreased activities of proteases. Thus, a more effective virus is shed through the respiratory tract (Lowen et al., 2007; Lowen and Steel, 2014).

The second mechanism is the enhanced survivability of the coronaviruses on surfaces at lower temperatures (Casanova et al., 2010; Chan et al., 2011; van Doremalen, Bushmaker, and Munster, 2013). Direct contact with contaminated surfaces is one of the main routes of coronavirus transmission. Previous research has shown that high temperatures can rapidly inactivate coronaviruses (Lai, Cheng, and Lim, 2005; Pirtle and Beran, 1991). The inactivation on surfaces at high temperatures can be attributed to two different means, depending on the relative humidity (Casanova et al., 2010). At low levels of relative humidity, when the temperature increases the virus is desiccated. The desiccation process results in lipid membrane phase changes, oxidation, and Maillard reactions (Cox, 1993). Consequently, the virus is inactivated. At high levels of relative humidity, inactivation with high temperature occurs mainly as a result of the accumulation of viral capsids at the air-water interface and the resultant structural damages to the virus (Thompson, Flury, Yates, and Jury, 1998). When the relative humidity is moderate, both processes occur and result in the inactivation of coronavirus on surfaces (Casanova et al., 2010).

Several laboratory studies have been conducted on the effects of temperature on transmission and survival of prior coronaviruses such as severe acute respiratory syndrome (SARS) and the Middle East respiratory syndrome (MERS). For instance, (Casanova et al., 2010) used two surrogate viruses with similar transmissibility potential to the SARS coronavirus to model the effects of air temperature on the coronavirus survival on surfaces. The authors demonstrated a negative effect of air temperature on virus survival on steel surfaces. Similarly, (Chan et al., 2011) conducted a laboratory study on the survival of SARS coronavirus as a function of temperature. They found that high temperatures led to a rapid decrease in the virus’s survival on surfaces. Apart from these laboratory experiments, several field studies have been conducted on the effect of temperature, along with other environmental factors, on the spread rate of SARS disease during the SARS epidemic of the year 2003 (Tan et al., 2005; Yuan et al., 2006). For example, (Lin, Fong, Zhu, and Karlberg, 2006) found that SARS incidences increased by a factor of 18.18 in days with a lower temperature compared with days with a high temperature in Hong Kong during March to May of 2003.

Since the start of the novel coronavirus outbreak, there have been few empirical studies looking at the role of temperature on the incident rate of COVID-19 cases. In one of the first studies on the subject, (Wang, Tang, Feng, and Lv, 2020) constructed a measure of daily effective reproductive number as a proxy for the transmission severity of COVID-19. The authors then used the measure in order to test the effects of temperature and relative humidity on this proxy. Their results using data on 100 infected cities in China indicated a significant decrease in the transmission severity of COVID-19 with an increase in temperature. Similarly, using data on confirmed COVID-19 cases in 188 states or provinces across the world as of February 29, (Bannister-Tyrrell, Meyer, Faverjon, and Cameron, 2020) have shown a negative relationship between average temperature and number of confirmed cases of COVID-19. Similar results have been found with data on confirmed cases in China (Xie and Zhu, 2020). Consistent with this notion, researchers have found that areas with significant community transmission of COVID-19, such as Northern Italy, Iran, and South Korea, had a similar average temperature of 5-11 degree Celsius, in 20-30 days prior to the first community spread death (Sajadi et al., 2020). As another example, scholars used COVID-19 mortality data of Wuhan, China for the period between January 20 to February 29 and found a significant negative relationship between the number of COVID-19 related deaths in Wuhan and the city’s ambient temperature (Li et al., 2020).

The above discussion points to a negative relationship between temperature and the survival and transmission of COVID-19. In testing this relationship, we make several contributions. Most importantly, the results of this study can help in planning for the successful management of COVID-19 spread in the future. Also, we contribute to the literature on the implications of climate on critical social phenomena. This line of research has shown positive effects of temperature on criminal activities (Ranson, 2014), negative effects of exposure to extreme temperatures on birth weight (Deschênes, Greenstone, and Guryan, 2009), and negative effects of global warming on home prices (Butsic, Hanak, and Valletta, 2011).

## 3. Data and Sample Selection Strategy

The daily county-level data on confirmed cases of Covid-19 is extracted from the Centers for Disease Control and Prevention (CDC). I use the county-level population estimates in 2019 from (SEER, 2019) to calculate the daily new confirmed cases per 100,000 residents. The daily temperature data is from the Global Summary of the Day data files produced by the National Oceanic and Atmospheric Administration (NOAA). It gathers a summary of weather data, including surface temperature, across the globe in a daily basis. Within the US boundaries, there are over 2,500 stations located across 1,600 counties. We compute the county temperature by averaging the data of reporting stations within each county. We impute the values for counties with missing data by taking the average of all neighboring counties’ temperature for a given day. The Covid-19 data is then merged with the temperature data. We drop counties for which the population density was above three standard deviations from the mean of population density in 2019. The main reason for this sample restriction is that the new pandemic is contagious in short distances. Not only it is harder for residents of this county to keep the effective distance but also the knowledge of the effective distance was unknown for weeks. Therefore, the effect of temperature on the spread of the pandemic is potentially distorted among these counties. However, the results are robust without this sample restriction. Figure 1 shows the geographic distribution of the long-difference of new cases (top panel) and temperature (bottom panel) across US counties.

**Figure 1.**
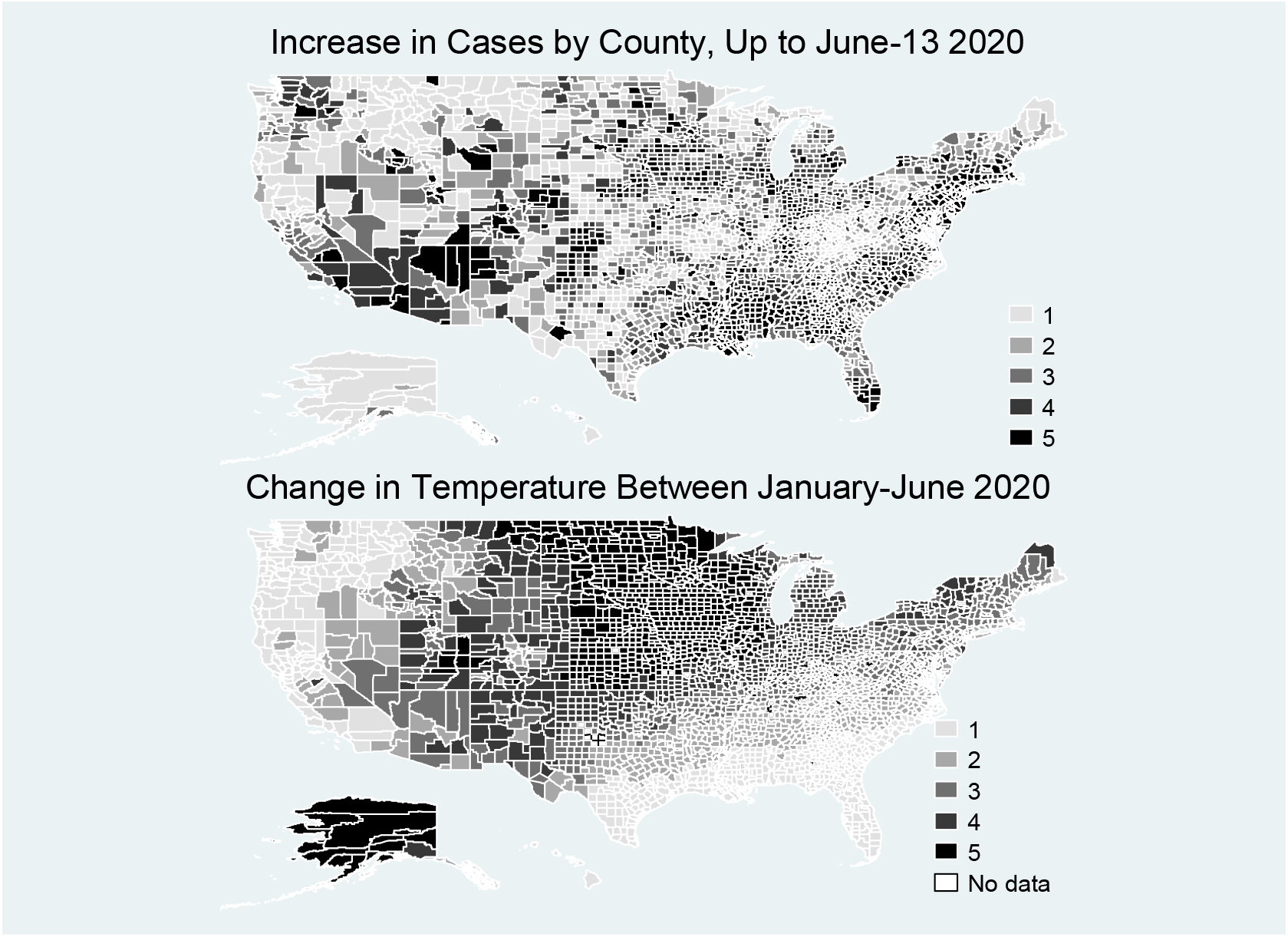
Changes in Temperature and Confirmed Cases between 22-January and 13-June 2020 Across US Counties.

The final sample includes 3,048 counties that cover all days between January 22, 2020 to June 13, 2020. A summary statistic of this sample is reported in Table 1. On average, 2.63 new confirmed cases occur in each county-day. Over the sample period, the average temperature was about 51 °*F*.

**Table 1.** Summary Statistics.

|  | Mean | Median | SD |
| --- | --- | --- | --- |
| New Cases per 100,000 Population | 2.63 | 0.00 | 21.24 |
| Average Temperature (F) | 51.59 | 51.77 | 16.30 |
| ***County Characteristics at 2018:*** |  |  |  |
| Population | 99,465.10 | 25642.00 | 324,353.01 |
| Population Density | 155.82 | 41.89 | 377.44 |
| % Whites | 85.98 | 92.88 | 16.14 |
| % Blacks | 9.79 | 3.02 | 14.52 |
| % Male | 50.11 | 49.69 | 2.24 |
| % Aged 15-30 | 18.64 | 18.10 | 3.96 |
| % Aged 30-50 | 23.29 | 23.29 | 2.59 |
| % Foreign Born | 4.24 | 2.01 | 5.36 |
| Unemployment Rate (%) | 4.58 | 4.30 | 1.62 |
| Personal Income Per Capita (\$) | 41,691.07 | 39,809.78 | 11,146.50 |
| Observation |  | 438,314 |  |

## 4. Empirical Strategy

The main source of identification is the plausibly exogenous changes in temperature across counties and over days. The identification strategy compares the new confirmed cases in counties that have higher within-week raises in temperature to those counties with lower within-week increases in temperature. It lies on the assumption that in the absence of any changes in temperature, the spread of the novel Coronavirus in counties with higher changes in temperature follows the same path and is influenced by the same factors as its spread in counties with lower changes in temperature.

There are a series of confounding factors that may bias the coefficients. We attempt to control for these observables and unobservables using a wide array of fixed effects. Each county has some characteristics that potentially affect its vulnerability and the speed of the spread of pandemics. For instance, population density, the share of foreign-born residents, or the composition of local industries, and occupational types are definitely correlated with exposure to the new disease. As long as these characteristics do not change with time, county fixed effect controls for them. Time fixed effects account for all unobservable features common across counties such as the spread of knowledge about social distancing. However, there are unobservable factors that are specific to a county that also change over time. For example, new kits for screening arrives which increases testing capacity and raises the number of confirmed cases. To account for these potentially confounding factors, we can also control for a series of the county by week fixed effects.

The empirical strategy can be summarized in the Ordinary-Least-Square regressions of the following form:

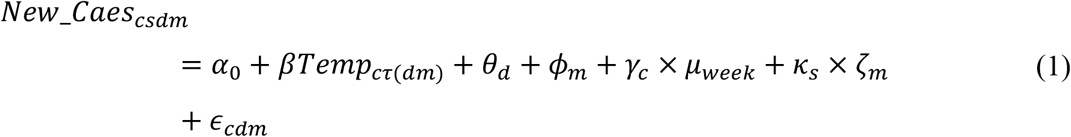

Where *New*_*Case* is the new confirmed cases in county *c* and day *d* of month *m*. The variable *Temp* is the temperature of the county in *τ* periods before. The choice of how many days before the new case is confirmed depends on the incubation period of Covid-19. The exact period between exposure and infection is still to be known. However, some reports of the World Health Organization suggest that the average incubation period is 5-6 days while it can be as long as 14 days (WHO, 2020). (Lauer et al., 2020) also report a median of 5 days. In our preferred specification we focus on the average temperature in the last three to five days, three to eight days, and also the average of the last three to fourteen.

The parameters *θ* and *ϕ* are a series of dummies to capture the main effects of day and month. In this model, the main effects of the county, *γ*_*c*_, are allowed to vary by week. To account for time trend features that are specific to each state, such as stay-at-home mandates, that could vary by month we add an interaction between state fixed effects, *κ*, with month fixed effects, *ζ*. We also add a series of state-specific time (day-month) trend in some specifications. Finally, *ϵ* is a disturbance term. Since the spread of the new pandemic depends on the social interaction of local residents, the error terms are possibly serially correlated. We cluster the standard errors at the county level to account for this serial correlation in the error terms.

## 5. Results

### 5.1. Main Results

The main results are reported in Table 2. An increase of one degree^6^ in temperature during the last 3-14 days is associated with a reduction of 0.048 cases per 100,000 county population. Adding state-by-month fixed effects (column 2) and also a state-specific linear trend (column 3) changes the magnitude of the coefficients only slightly. An increase of 16.3 degrees in temperature, the standard deviation of temperature over the sample period, reduces the new cases by 0.75 incidences per day. This is equivalent to an approximately 28% reduction from the mean of new confirmed cases over the sample. The respective coefficients are economically large and statistically significant at 1%. Assuming an incubation period of 3-8 days, the coefficient in our full model (column 3) shows a significant relationship at 5% between the average temperature during the incubation period and the number of new cases per 100,000 population. Specifically, with every one-degree increase in the average temperature, the number of new cases decreases by 0.018. This result has practical significance as it shows that new confirmed cases decrease by 0.29, which corresponds to a reduction of 11% from the mean when the average temperature increases by one standard deviation. Columns 1 and 2 show that the effect is quite robust if we exclude the state-by-month fixed effects and state-specific linear time trends.

**Table 2.** Main Results: The Effect of Temperature on Prevalence of New Confirmed Cases for Covid19.

|  | *Outcome: New Cases per 100,000 Population* |  |  |
| --- | --- | --- | --- |
|  | Model (1) | Model (2) | Model (3) |
| *Average Temperature in X Prior Days:* |  |  |  |
| $X = [3,14]$ | -0.0484***<br>(0.0127) | -0.0452***<br>(0.0146) | -0.0457***<br>(0.0146) |
| $X = [3,8]$ | -0.0180**<br>(0.0079) | -0.0177**<br>(0.0078) | -0.0179**<br>(0.0078) |
| $X = [3,5]$ | -0.0136***<br>(0.0052) | -0.0140***<br>(0.0047) | -0.0145***<br>(0.0047) |
| Day Dummies | Yes | Yes | Yes |
| Month Dummies | Yes | Yes | Yes |
| County-by-Week Fixed Effects | Yes | Yes | Yes |
| State-by-Month Fixed Effects | No | Yes | Yes |
| State-Specific Day-by-Month Trend | No | No | Yes |
| Observations | 438,314 | 438,314 | 438,314 |

Notes. Each cell represents a separate regression. Standard Errors, reported in parentheses, are clustered at the county level.
\*Significant at the 10% level.
\*\*Significant at the 5% level.
\*\*\*Significant at the 1% level.

The average US temperature varies from roughly 68 degrees in May (the end of our sample) to around 80 degrees in its peak in August. If we assume that the incubation is between 3-14 days, the average rise in temperature can reduce the new cases by 0.54 fewer incidences per 100,000 county population per day. To put it into perspective, this rise in temperature avoids roughly 1,800 new cases per day in the country.

### 5.2. Robustness Checks

These results could be driven by a sub-population in the data and be smaller or even opposite-signed in other sub-populations. If so, one can detect the primarily impacted population and revise any policy suggestion henceforth. Therefore, we checked the robustness of our results by creating several sub-samples which can theoretically affect our estimates. We ran our analysis on each of these sub-samples, as depicted in Table 3. We used the same model and the same dependent variable as in our main analysis with average temperature during the three assumed incubation periods (the last 3-5, 3-8, and 3-14 days) as the independent variables. In all regressions, a full specification of equation 1 is applied where in addition to fixed effects, state-by-month interaction and state-time linear trend is used. As can be seen in Table 3, when we exclude the six highly infected states, the effects on temperature on new cases remain statistically significant at 1% and the coefficients change slightly. Next, we divided our sample based on unemployment rate, percentage of male in the county population, percentage of residents with the age 15-30, population density, and percentage of foreign-born residents. Our analyses show that the association between the average temperature and the new cases per 100,1000 population is robust in a majority of sub-samples. However, the relationship goes away for the sub-sample of counties above the median population of 15-30 years old and above the median population density.

**Table 3.** Robustness of The Main Results by Different Subsamples.

| *Subsamples:* | *Outcome:* |  |  |
| --- | --- | --- | --- |
|  | Average<br>Temperature in<br>The Past = [3,5]<br>Days | Average<br>Temperature in<br>The Past = [3,8]<br>Days | Average<br>Temperature in<br>The Past =<br>[3,14] Days |
| Excluding New York, New Jersey, California,<br>Pennsylvania, Michigan, and Massachusetts | -0.0162***<br>(0.0051) | -0.0175**<br>(0.0084) | -0.0418***<br>(0.0157) |
| Below Median Unemployment Rate | -0.0071*<br>(0.0043) | -0.0170***<br>(0.0053) | -0.0393**<br>(0.0184) |
| Above Median Unemployment Rate | -0.0241***<br>(0.0079) | -0.0205<br>(0.0139) | -0.0408**<br>(0.0216) |
| Below Median Share of Male | -0.0104***<br>(0.0034) | -0.0170***<br>(0.0053) | -0.0410***<br>(0.0011) |
| Above Median Share of Male | -0.0202**<br>(0.0090) | -0.0205<br>(0.0139) | -0.0556***<br>(0.0214) |
| Below Median of Share of 15-30 Aged Residents | -0.0115***<br>(0.0038) | -0.0201***<br>(0.0052) | -0.0503***<br>(0.0105) |
| Above Median of Share of 15-30 Aged Residents | -0.0163**<br>(0.0078) | -0.0123<br>(0.0137) | -0.0331<br>(0.0283) |
| Below Median Population Density | -0.0194**<br>(0.0083) | -0.0318**<br>(0.0125) | -0.0745***<br>(0.0171) |
| Above Median Population Density | -0.0103**<br>(0.0048) | -0.0042<br>(0.0098) | -0.0182<br>(0.0211) |
| Below Median of Share of Foreign-Born<br>Residents | -0.0183***<br>(0.0070) | -0.0285***<br>(0.0105) | -0.0492***<br>(0.0148) |
| Above Median of Share of Foreign-Born<br>Residents | -0.0098*<br>(0.0052) | -0.0046<br>(0.0107) | -0.0462**<br>(0.0230) |

Notes. Each cell represents a separate regression. The independent variable is new cases per 100,000 county population. All regressions include a series of dummies for day and month, state-specific day-month trend, and county-by-week fixed effects. Standard Errors, reported in parentheses, are clustered at the county level.
\*Significant at the 10% level.
\*\*Significant at the 5% level.
\*\*\*Significant at the 1% level.

As a further analysis, we also apply a dynamic GMM model with a series of fixed effects. A summary of our dynamic model is reported in Table 4. Although the point estimates are slightly smaller than those of the fixed-effect model, they are quite comparable, similar in sign, and significant at 1% level. For instance, using an incubation period between 3-14 days and applying a two-step GMM model, 10 degrees rise in temperature is associated with 0.53 fewer incidence of new confirmed cases per 100,000 population, equivalent to a 20% reduction from the mean of new cases at the county level.

**Table 4.** Robustness of The Main Results Using Dynamic GMM.

|  | *Outcome: New Cases per 100,000 Population* |  |  |  |
| --- | --- | --- | --- | --- |
|  | One-Step GMM | Two-Step GMM | One-Step GMM | Two-Step GMM |
|  | (1) | (2) | (3) | (4) |
| Average Temperature in The Past [3,5] Days | -0.0322***<br>(0.0057) | -0.0322***<br>(0.0001) | -0.0159***<br>(0.0058) | -0.0159***<br>(0.0001) |
| Average Temperature in The Past [3,8] Days | -0.0626***<br>(0.0084) | -0.0626***<br>(0.0001) | -0.0426***<br>(0.0087) | -0.0425***<br>(0.0001) |
| Average Temperature in The Past [3,14] Days | -0.1095***<br>(0.0155) | -0.1095***<br>(0.00002) | -0.0528***<br>(0.0166) | -0.0527***<br>(0.00016) |
| Day and Month Fixed Effects | Yes | Yes | Yes | Yes |
| County Fixed Effects | Yes | Yes | Yes | Yes |
| State-by-Month Fixed Effects | No | No | Yes | Yes |
| Observations | 438,314 | 438,314 | 438,314 | 438,314 |

Notes. Each cell represents a separate regression. Sargan test of overidentification restriction is rejected in all regressions and two-step GMM is the preferred model.
\*Significant at the 10% level.
\*\*Significant at the 5% level.
\*\*\*Significant at the 1% level.

## 6. Discussion and Conclusion

The recent pandemic (COVID-19) is considered as the most crucial global health challenge that impacted both developing and developed countries and their economies since World War II. The U.S. has one-third of the world’s COVID-19 confirmed cases so far and has the largest number of death due to the pandemic across countries. This study suggested that daily temperature influences the daily COVID-19 confirmed new cases in the U.S. at the county-level. we employed panel data from January 22, 2020 to June 13, 2020 covering all U.S. counties for confirmed cases of the new pandemic as well as the temperature. We used various static and dynamic models to measure the impact of daily temperature on the number of COVID-19 confirmed cases at the county level. The main static model indicates that an increase of one degree in temperature during the last 3-14 days is associated with a reduction of 0.045 cases per 100,000 population at the county-level. The dynamic results are consistent with the static findings and confirm that the higher temperature in a county most likely will drop the number of COVID-19 confirmed cases. The results indicate that the two-step system GMM estimator is more appropriate than a one-step system GMM estimator for measuring the impact of temperature on the number of cases. These findings could be used by state or national policymakers to optimize their decisions regarding new policies, regulations, and budget allocation.

## Data Availability

The data is available upon request

## Footnotes

4 Prices of agricultural commodities have been fallen by 20% as demand from hotels and restaurants has been drop (Jayashree Bhosale, 2020; Nicola et al., 2020).

5 This is the minimum amount that Fed can determine for the interest rate.

6 The temperature units are all in Fahrenheit degree.

